# Frailty Index Comparison in Predicting Postoperative Outcomes in Hepatic Cystic Echinococcosis: A Nested Case-Control Study

**DOI:** 10.1101/2025.08.01.25332747

**Authors:** Carlos Manterola, Josue Rivadeneira, Luis Alvarado, Luis Grande

**Affiliations:** Department of Surgery, Center of Morphological and Surgical Studies (CEMyQ), Universidad de La Frontera, Temuco, Chile; PhD Program in Medical Sciences, Universidad de La Frontera, Temuco, Chile; Zero Biomedical Research, Quito, Ecuador; Faculty of Medicine, Universidad Central del Ecuador, Quito, Ecuador; Department of Surgery, Universitat Autònoma de Barcelona, Barcelona, Spain; Hospital del Mar Research Institute (IMIM), Barcelona, Spain; Reial Academia de Medicina de Catalunya, Barcelona, Spain

**Author notes:** **Corresponding author**: Dr. Carlos Manterola. Universidad de La Frontera. Manuel Montt 112, office 408, Temuco, Chile. **Author’s contribution to the manuscript.** <u>Conception and design</u>: CM, JR and LG. <u>Analysis and interpretation</u>: CM, JR, and LG. <u>Data collection</u>: CM, JR, and LA. <u>Writing the article</u>, critical revision of the article: CM, JR, LA and LG. <u>Obtaining funding</u>: CM.

**Keywords:** "Echinococcosis, Hepatic"[Mesh], "Frailty"[Mesh], "Frailty/surgery"[Mesh], "Postoperative Complications"[Mesh], "Case-Control Studies"[Mesh], "Prognosis"[Mesh]

## Abstract

**Background:** Postoperative complications (POC) following surgery for hepatic cystic echinococcosis (HCE) remains a persistent clinical challenge. The utility of frailty indices (FIs) as predictors of POC in this context has not been investigated. This study aimed to assess predictive value of three FIs—mFI-11, FRAIL, and PRISMA-7—for POC in patients undergoing surgery for HCE.

**Methodology/Principal findings:** Nested case–control study. Patients who underwent elective surgery for HCE (2012-2020), matched in a 1:1 case-to-control ratio were included. Cases were defined as patients with mFI-11 score ≥0.27, PRISMA-7 ≥3, or FRAIL ≥3; controls were those with mFI-11 <0.27, PRISMA-7 <3, or FRAIL <3. The primary outcome was POC. Surgical procedures included pericystectomy and hepatic resection. Descriptive statistics and bivariate analyses were applied. Logistic regression, odds ratios (OR) with 95% confidence intervals were calculated. 140 patients were included (70 cases and 70 controls). In multivariable logistic regression, mFI-11 ≥0.27 was identified as an independent prognostic factor for overall and severe POC (p<0.001 and p=0.02, respectively). On the other hand, PRISMA-7 ≥3 was also identified as an independent prognostic factor for overall and major POC (p=0.02 and p=0.03, respectively); and FRAIL scale an independent prognostic factor for overall POC (p=0.005).

**Conclusion/Significance:** Frail patients exhibited higher frequency and severity of POC compared to non-frail patients. mFI-11 demonstrated the best performance in predicting overall and severe POC.

**Author summary:** Cystic echinococcosis is a neglected zoonotic tropical disease that primarily affects impoverished pastoral communities worldwide. It involves a parasitic cycle between farm dogs and livestock, often linked to livestock farming practices and the feeding of infected offal to dogs. In humans, the disease causes significant morbidity and mortality through the development of cysts in various organs, most commonly the liver

Three frailty indices were applied to patients who underwent surgery for HCE, to assess their predictive capacity for POC development between frails (cases) and non-frails (controls). An mFI-11 ≥0.27 is an independent predictive factor for POC.

The importance of this finding lies in knowing that likelihood for overall POC in these patients with an mFI-11 ≥0.27 is five times higher.

## INTRODUCTION

Cystic echinococcosis, caused by the metacestode stage of *Echinococcus granulosus*, is among the parasitic zoonoses with the highest global burden on both human and animal health [1]. According to the World Health Organization (WHO), is globally distributed, with incidence rates reaching up to 50 cases per 100,000 population in endemic regions [2]. In Chile, which is considered an endemic country, the estimated national incidence rate is approximately 2 cases per 100,000 population. However, in certain regions such as La Araucanía, the incidence increases to 6 cases per 100,000 population [3].

Surgery remains the most effective therapeutic option for hepatic cystic echinococcosis (HCE), despite the variability in surgical techniques and approaches [4]. Although HCE is a benign disease, postoperative complications (POC) remain a significant unresolved concern. Reports from 2018 indicate complication rates as high as 63% [4-10], with 30-day mortality (30-DM) reaching up to 2% [8,11].

Despite the extensive literature addressing preoperative frailty, no study to date has evaluated the utility of frailty indices (FI) in predicting outcomes in patients undergoing elective surgery for HCE Frailty represents a state of increased vulnerability to stressors such as illness or trauma, leading to an impaired homeostatic response and a higher risk of complications and adverse outcomes. Although most associated with aging, frailty is not exclusive to older adults. It is a complex and multidimensional condition involving multiple organs and systems and may be associated with specific frailty phenotypes [12]. The underlying pathophysiology is thought to result from cumulative deficits in physical, psychosocial, or biological reserves. Factors, such as, physical deterioration, malnutrition, chronic diseases, and disability accelerate physiological aging, contributing to sarcopenia, decrease muscular strength, and chronic inflammation [12,13].

To assess and quantify frailty, more than 20 validated tools are available. While these instruments share certain characteristics, there is currently no universally accepted gold-standard. This diversity allows for flexibility in selecting the most appropriate assessment tool based on type of surgery, patient characteristics, and available resources [14]. Common instruments include modified Frailty Index (mFI), particularly its mFI-11 version [15-18], FRAIL scale [19], and PRISMA-7 questionnaire [20].

The aim of this study was to evaluate the predictive value of three frailty assessment tools (mFI-11, FRAIL scale, and PRISMA-7 questionnaire), for POC in patients with HCE undergoing elective surgical treatment.

## METHODS

This manuscript was prepared according to STROBE statement for observational studies reporting [21].

### Design

Nested case-control study in a concurrent cohort.

### Setting

The study was performed at Surgery Department, Universidad de La Frontera (Temuco, Chile).

### Participants

Consecutive patients who underwent elective open surgery for HCE between 2012 and 2020 with a minimum follow-up of 4 years were included. Patients who underwent laparoscopic surgery were excluded. Cases were defined as patients who underwent elective surgery for HCE with a score above the cut-off point on any of the three FI under study (mFI-11 ≥ 0.27, PRISMA-7 ≥ 3, or FRAIL ≥ 3). Cases and controls were matched in a 1:1 ratio based on age, sex, main cyst diameter, history of prior HCE surgery, ultrasonographic characteristics, and main cyst location (Figure 1).

**Figure 1.**
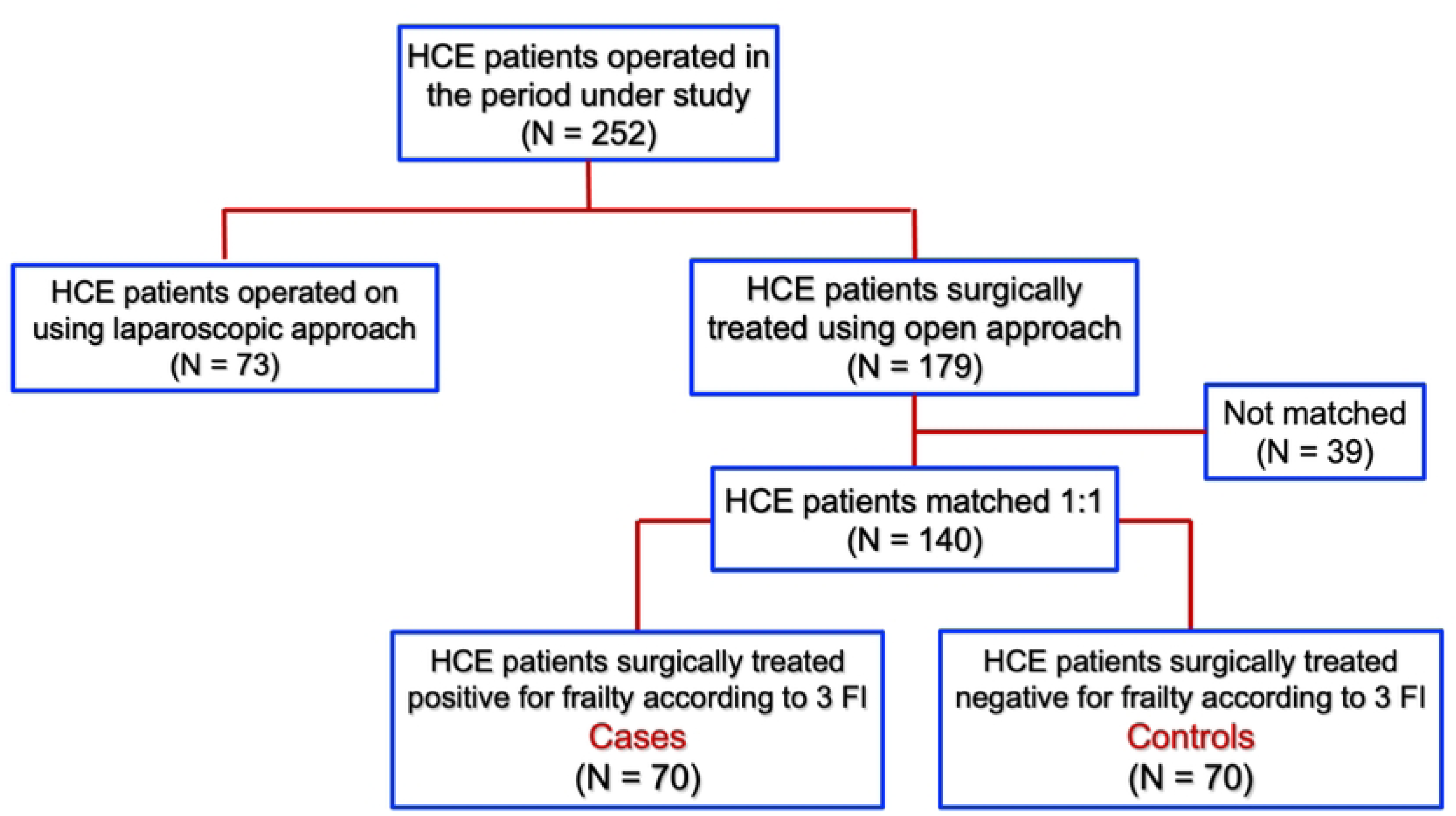
Flow chart of the study participants.

### Study Outcomes

Primary outcome was the occurrence of POC, defined as pathologic processes that affect patients after a surgical procedure, that may or may not be related to the disease for which the surgery was done, and they may or may not be direct results of the surgery (https://www.ncbi.nlm.nih.gov/mesh/68011183); graded using the Clavien-Dindo classification [22]. Severe complications were classified as those corresponding to Clavien-Dindo grade >IIIa. This was measured as a binary variable (yes/no). Secondary outcomes included 30-DM (number of days), need for reintervention (yes/no), and disease recurrence (yes/no), defined as any detectable abdominal cyst during >48 month follow-up.

### Additional variables of interest

age, sex; preoperative variables (simultaneous cysts, previous surgery for HCE, ultrasonography classification [23], number of hepatic cysts, location of principal cyst), intraoperative variables (biliary communication, type of cyst surgery, residual cavity treatment), postoperative characteristics; and HCE evolutionary complications: immunoallergic [24], intraperitoneal rupture [25], intrabiliary rupture [26], cholangiohydatidosis with or without acute cholangitis [27], cyst infection with abscess formation [28], and hepatothoracic transit [29]. American Society of Anesthesiologists (ASA), score grouping patients into I-II and III-IV; operating time, and length of hospital stance (LHS) were also measured.

### Data sources/measurements

mFI-11 assigns one point for each of 11 potential frailty components, resulting in a score from 0 to 11 [16], and patients were classified as cases when they had ≥3 deficits (mFI ≥0.27), and controls when they had ≤2 deficits (mFI <0.27). FRAIL scale ranges from 0 to 5 [19]; and patients were classified as cases when they had <u>≥</u> 3 points, and controls when they had ≤ 2 points. PRISMA-7 assigns 1 point for each of 7 potential frailty components [20]. Patients were classified as cases when they had ≥ 3points, and controls when they had ≤ 2 points.

### Surgical technique

All patients, cases and controls, underwent open surgery, including evacuation of the cystic content, endocystectomy, and suturing of biliary leaks. Procedures included total/subtotal pericystectomy, and hepatic resection, cystectomy or unroofing were excluded. Selected cases received external drainage. Albendazole was administered postoperatively for 3 months in patients with previous HCE surgery or active/transitional cysts.

### Bias

Multiple strategies were implemented to reduce potential sources of bias: a) minimum follow-up period of 48 months for all participants; b) controls were selected from the same population as cases to ensure similar exposure; c) a simple random sampling method was applied to select controls; d) matching was performed on key variables to reduce potential confounders; and e) data collection was conducted by investigators who did not participate in the surgical procedures (JR, LA).

### Sample size

Calculation was based on POC rates of 28% for cases and 11% for controls, assuming a type I error rate of 5% and a statistical power of 80%, requiring a minimum of 66 cases and 66 controls [31].

### Statistical Analysis

Data were extracted from CEMyQ database. After exploratory analysis, descriptive statistics and bivariate test (t-tests, Pearson’s Chi-square, Fisher’s exact tests with Savalei’s correction for zero-cells frequencies [32]) were performed. Logistic regression was used to estimate odds ratios (OR) and their respective 95% confidence intervals (95% CI), and predictive performance was evaluated using area under the receiver operating characteristics curve (AUC), and its comparison applying DeLong test [33]. Analyses were conducted using Stata 11.0 statistical package

### Ethics

The study was conducted in accordance with the Declaration of Helsinki [34], and was approved by the Institutional Review Board (CEISH-UFRO, protocol code 007_25). Written consent was obtained from all participants.

## RESULTS

Out of 252 surgically treated HCE patients, 140 were matched and included in the final analysis (70 cases and 70 controls) (Figure 1).

Table 1 displays continuous variables, with significant differences observed in operating time, and scores for mFI-11, FRAIL, and PRISMA-7.

**Table 1.**
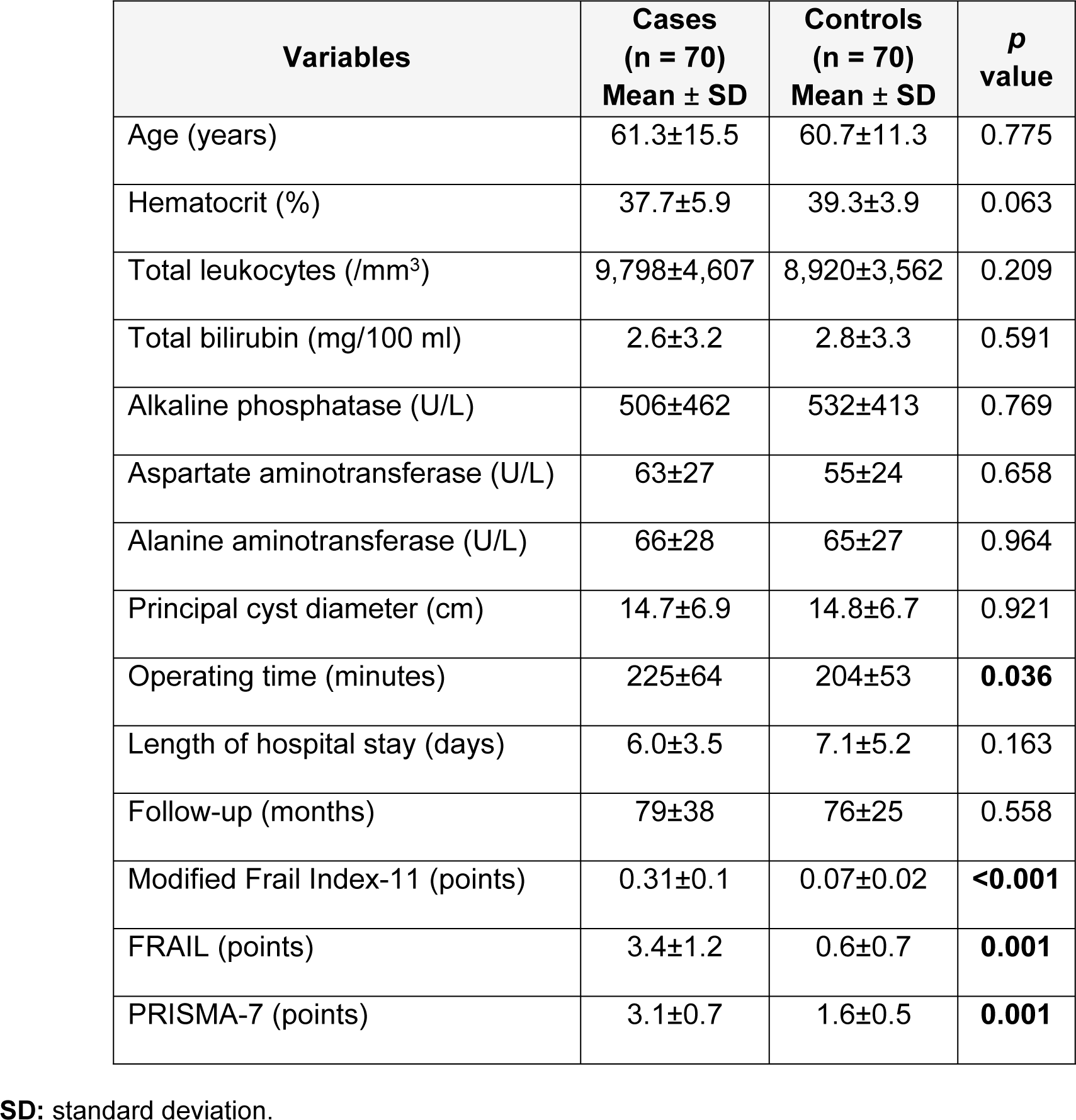
Distribution of clinical variables between cases and controls.

| Variables | Cases<br>(n = 70)<br>Mean ± SD | Controls<br>(n = 70)<br>Mean ± SD | p<br>value |
| --- | --- | --- | --- |
| Age (years) | 61.3±15.5 | 60.7±11.3 | 0.775 |
| Hematocrit (%) | 37.7±5.9 | 39.3±3.9 | 0.063 |
| Total leukocytes (/mm<sup>3</sup>) | 9,798±4,607 | 8,920±3,562 | 0.209 |
| Total bilirubin (mg/100 ml) | 2.6±3.2 | 2.8±3.3 | 0.591 |
| Alkaline phosphatase (U/L) | 506±462 | 532±413 | 0.769 |
| Aspartate aminotransferase (U/L) | 63±27 | 55±24 | 0.658 |
| Alanine aminotransferase (U/L) | 66±28 | 65±27 | 0.964 |
| Principal cyst diameter (cm) | 14.7±6.9 | 14.8±6.7 | 0.921 |
| Operating time (minutes) | 225±64 | 204±53 | **0.036** |
| Length of hospital stay (days) | 6.0±3.5 | 7.1±5.2 | 0.163 |
| Follow-up (months) | 79±38 | 76±25 | 0.558 |
| Modified Frail Index-11 (points) | 0.31±0.1 | 0.07±0.02 | **<0.001** |
| FRAIL (points) | 3.4±1.2 | 0.6±0.7 | **0.001** |
| PRISMA-7 (points) | 3.1±0.7 | 1.6±0.5 | **0.001** |

**SD:** standard deviation.

On the other hand, table 2 shows preoperative variables, revealing significant differences in the incidence of postoperative complications (OR: 2.5; p=0.005), and ASA (OR: 0.11; p<0.001). No significant differences were found in ultrasonographic features, number of cysts, biliary communications, type of surgery, or residual cavity treatment.

**Table 2.** Distribution of perioperative variables between cases and controls.

| Variables | Cases<br>(n = 70)<br>n (%) | Controls<br>(n = 70)<br>n (%) | OR | 95% CI | p<br>value |
| --- | --- | --- | --- | --- | --- |
| Sex |  |  |  |  |  |
| Female | 32 (45.7) | 32 (45.7) | 0.8 | 0.5-2.0 | 1.000 |
| Male | 38 (54.3) | 38 (54.3) |  |  |  |
| Simultaneous cysts* |  |  |  |  |  |
| Yes | 22 (31.4) | 30 (42.9) | 0.6 | 0.3-1.2 | 0.107 |
| No | 48 (68.6) | 40 (57.1) |  |  |  |
| Evolutionary complications of HCE |  |  |  |  |  |
| Yes | 34 (48.6) | 19 (27.1) | **2.5** | **1.3-5.1** | **0.005** |
| No | 36 (51.4) | 51 (72.9) |  |  |  |
| Previous surgery for HCE |  |  |  |  |  |
| Yes | 10 (14.3) | 11 (15.7) | 0.9 | 0.4-2.3 | 0.814 |
| No | 60 (85.7) | 59 (84.3) |  |  |  |
| ASA |  |  |  |  |  |
| I or II | 35 (50.0) | 63 (90.0) | **0.1** | **0.05-0.3** | **<0.001** |
| III or IV | 35 (50.0) | 7 (10.0) |  |  |  |
| Ultrasonography |  |  |  |  |  |
| CE 1 | 23 (32.9) | 28 (40.0) | --- | --- | 0.848 |
| CE 2 | 14 (19.9) | 12 (17.1) |  |  |  |
| CE 3a | 17 (24.3) | 16 (22.9) |  |  |  |
| CE 3b | 16 (22.9) | 14 (20.0) |  |  |  |
| Number of hepatic cysts |  |  |  |  |  |
| 1 | 47 (67.1) | 42 (60.0) | --- | --- | 0.091 |
| 2 | 10 (14.3) | 20 (28.6) |  |  |  |
| ≥3 | 13 (18.6) | 8 (11.4) |  |  |  |
| Location of principal cyst |  |  |  |  |  |
| Right lobe | 41 (58.6) | 42 (60.0) | --- | --- | 0.974 |
| Left lobe | 13 (18.6) | 12 (17.1) |  |  |  |
| Central or bilateral | 16 (22.9) | 16 (22.9) |  |  |  |
| Biliary communication |  |  |  |  |  |
| 0 | 29 (41.2) | 23 (32.9) | --- | --- | 0.204 |
| 1 | 28 (40.2) | 25 (35.7) |  |  |  |
| ≥2 | 13 (18.6) | 22 (31.4) |  |  |  |
| Principal cyst surgery |  |  |  |  |  |
| Partial pericystectomy | 42 (60.0) | 49 (70.0) | --- | --- | 0.462 |
| Total pericystectomy | 17 (24.3) | 13 (18.6) |  |  |  |
| Hepatic resections | 11 (15.7) | 8 (11.4) |  |  |  |
| External biliary drainage |  |  |  |  |  |
| Yes | 19 (27.1) | 17 (24.3) | 1.1 | 0.7-1.6 | 0.699 |
| No | 51 (72.9) | 53 (75.7) |  |  |  |
| Residual cavity treatment |  |  |  |  |  |
| Capitonnage | 17 (24.3) | 20 (28.6) | --- | --- | 0.373 |
| Omentoplasty | 12 (17.1) | 17 (24.3) |  |  |  |
| Nothing | 41 (58.6) | 33 (47.1) |  |  |  |

**OR:** odds ratio; **95% CI:** 95% confidence interval; **HCE:** Hepatic cystic echinococcosis
\*Simultaneous cysts in other location than the liver

Postoperative variables are shown in Table 3, revealing higher frequencies of overall POC (OR: 2.9; p=0.005) and severe POC (OR: 1.8; p=0.029) in the frailty group. No statistical differences were noted in reintervention, 30-DM, or recurrence. Reintervention occurred in 3 cases (biliary leak, evisceration, and eventration) and in 4 controls (biliary leak, evisceration, and two incisional hernias). Two cases died with 30 days ( one from sepsis and other from pulmonary thromboembolism), with no deaths in the control group. Recurrence was documented in 2 controls (at 26 and 34 months of follow-up) within the cases, and in 1 case (36 months of follow-up) of the controls.

**Table 3.** Distribution of postoperative variables between cases and controls.

| Variables | Cases<br>(n = 70)<br>n (%) | Controls<br>(n = 70)<br>n (%) | OR | 95% CI | p<br>value |
| --- | --- | --- | --- | --- | --- |
| Overall POC |  |  |  |  |  |
| Yes | 23 (32.9) | 10 (14.3) | **2.9** | **1.3-6.8** | **0.005** |
| No | 47 (67.1) | 60 (85.7) |  |  |  |
| Severe POC (Clavien ≥ IIIa) | | | | | |
| Yes | 7 (10.0) | 1 (1.4) | **1.8** | **1.3-2.5** | **0.029** |
| No | 63 (90.0) | 69 (98.6) |  |  |  |
| Reoperations |  |  |  |  |  |
| Yes | 3 (4.3) | 4 (5.7) | 0.7 | 0.2-3.4 | 0.954 |
| No | 67 (95.7) | 66 (94.3) |  |  |  |
| 30-day mortality |  |  |  |  |  |
| Yes | 2 (2.9) | 0 (0.0) | 3.1 | 0.3-30.4 | 0.102 |
| No | 68 (97.1) | 70 (100.0) |  |  |  |
| Recurrence |  |  |  |  |  |
| Yes | 1 (1.4) | 2 (2.9) | 0.5 | 0.1-5.6 | 0.576 |
| No | 69 (98.6) | 68 (97.1) |  |  |  |

**OR:** odds ratio; **95% CI:** 95% confidence interval; **POC:** postoperative complications

Among the FI, mFI-11 demonstrated better performance for overall (OR: 4.8; p=0.0001) and severe POC (7.3; p=0.023). Patients with an mFI-11 ≥ 0.27 had nearly fivefold higher odds of any POC and sevenfold greater odds of severe POC. While FRAIL and PRISMA-7 also showed positive associations (ORs: 2–3), their wider 95% CIs limited statistical precision (Table 4).

Logistic regression adjusting for potential confounding variables (operating time, HCE evolutionary complications, and ASA) confirmed mFI-11 ≥ 0.27 as an independent prognostic factor for overall POC (OR 4.8; p=0.0001; AUC: 0.7624) and severe POC (OR 10.7; p=0.022; AUC: 0.7624). In addition, it was verified that PRISMA-7 ≥ 3 points is an independent prognostic factor for overall POC (OR 2.5; p=0.022; AUC: 0.6093) and severe POC (OR 10.3; p=0.026; AUC: 0.6212). On the other hand, FRAIL scale is an independent prognostic factor for overall POC (OR 2.9; p=0.005; AUC: 0.7389). See Table 4.

**Table 4.** Comparison of Surgical Outcomes between cases and controls according to applied frailty indexes.

| Outcomes | mFI-11 |  |  |  |  |  | FRAIL scale |  |  |  |  |  | PRISMA-7 |  |  |  |  |  |
| --- | --- | --- | --- | --- | --- | --- | --- | --- | --- | --- | --- | --- | --- | --- | --- | --- | --- | --- |
|  | Cases<br>(n=52) | Controls<br>(n=70) | OR | CI 95% | p<br>value | AUC | Cases<br>(n=70) | Controls<br>(n=70) | OR | CI 95% | p<br>value | AUC | Cases<br>(n=54) | Controls<br>(n=70) | OR | CI 95% | p<br>value | AUC |
| Overall POC, yes | 23 (44.2) | 10 (14.3) | **4.8** | **2.0-11.3** | **0.0001** | **0.762** | 23 (32.9) | 10 (14.3) | **2.9** | **1.3-6.8** | **0.005** | 0.739 | 16 (29.6) | 10 (14.3) | **2.5** | **1.1-6.1** | **0.022** | 0.609 |
| Severe POC, yes | 7 (13.5) | 1 (1.4) | **10.7** | 1.3-90.2 | **0.022** | **0.763** | 7 (10.0) | 1 (1.4) | 7.7 | 0.9-54.1 | 0.069 | **0.819** | 7 (13.0) | 1 (1.4) | **10.3** | 1.2-86.3 | **0.026** | 0.621 |
| Reoperation, yes | 3 (5.6) | 4 (5.7) | 1.0 | 0.2-4.7 | 0.989 | 0.549 | 3 (4.3) | 4 (5.7) | 0.7 | 0.2-3.4 | 0.954 | 0.568 | 1 (1.9) | 4 (5.7) | 0.3 | 0.1-2.9 | 0.204 | 0.660 |
| 30-day mortality, yes | 2 (3.7) | 0 (0.0) | 2.8 | 0.3-31.7 | 0.317 | 0.763 | 2 (2.9) | 0 (0.0) | 2.1 | 0.2-23.2 | 0.561 | **0.833** | 2 (3.7) | 0 (0.0) | 2.7 | 0.2-30.5 | 0.341 | **0.826** |
| Recurrence, yes | 1 (1.9) | 2 (2.9) | 0.7 | 0.1-7.6 | 0.742 | 0.619 | 1 (1.4) | 2 (2.9) | 0.5 | 0.1-5.6 | 0.577 | 0.543 | 0 (0.0) | 2 (2.9) | 0.6 | 0.1-7.1 | 0.986 | 0.804 |

**CI 95%:** confidence interval 95%; **POC:** postoperative complications; Severe **POC:** POC Clavien ≥ IIIa; **AUC:** area under the receiver operating characteristic curves

## DISCUSSION

### Summary of the evidence

Despite the high morbidity and mortality associated with surgery for HCE, to date, no studies have investigated the potential impact of frailty in this patient population. In this nested case-control study, we demonstrated not only that frail patients have a significantly higher incidence of overall and severe postoperative complications, but also that, among three frailty assessment tools, the mFI-11 showed the strongest predictive performance for both overall and severe POC.

In contexts other than HCE surgery, numerous studies have examined the relationship between frailty and perioperative outcomes. Although there is a general trend associating frailty with a higher rate of complications, the variability in findings is considerable [35-37]. This may be attributed to differences in study populations, research designs, and the frailty assessment tools applied [38,39]. Therefore, comparative evaluations across specific subpopulations are essential.

Although 30-DM was not significantly different, cases were more likely to present postoperative (48.6% vs. 27.1%; OR: 2.5; p=0.005) and had worse ASA (ASA I–II: 50% vs ASA III-IV: 90% OR: 0.1; p<0.001), suggesting a high baseline surgical risk.

No patients had extreme frailty scores, with maximum values of 6 (FRAIL), 4 (PRISMA) or 4 deficits (mFI-11). This may explain the weaker association between FI and severe POC or 30-DM.

Importantly, HCE is a benign condition often affecting young patients. In fact, 42.9% of all patients studied (48.6% of frails and 37.1% of non-frails), were aged 65 years or older. Moreover, despite scientific and technological advances, the high rates of postoperative complications (POC) associated with this condition have persisted over time, with little variation over the past 20 years. Therefore, this study illustrates that frailty is not exclusive to elderly or oncologic populations. These indices can be clinically useful even in younger cohorts with non-malignant conditions. Evidence on this point exists in other situations; for example, in cases of minimally invasive distal pancreatectomy and hepatectomies for benign disease [40-42]. Also, these findings may inform preoperative counseling and aid in identifying candidates for prehabilitation, particularly those in whom a higher risk of developing POC can be anticipated.

### Limitations

Limitations included external validity due to single-center, and internal validity due to a retrospective design and a small sample size. Nevertheless, the use of matched controls and regression adjustment strengthened internal validity and reduced potential confounding. Given its strong predictive value across surgical specialties, frailty assessment using mFI-11 offers a valuable tool for preoperative risk stratification [16,18].

## Conclusion

In conclusion, the mFI-11 index showed the strongest predictive performance for both overall and severe POC in patients underwent HCE.

## Financial Disclosure statement

This study was partially financed by the following grants:

DIUFRO DI23-0020. Dirección de Investigación, Universidad de La Frontera. **CM.**

The funders had no role in study design, data collection and analysis, decision to publish, or preparation of the manuscript.

## Data Availability Statement

All data are in the manuscript and/or supporting information files.

## Ethics Approval

The study protocol was approved by the Clinical Research Ethics Committee (CEIC) of Universidad de La Frontera (code 007_25).

## Statement

During the preparation of this work the authors not used IA tool/service.

## REFERENCES

1. Noguera LP, Charypkhan D, Hartnack S, Torgerson PR, Rüegg SR. The dual burden of animal and human zoonoses: A systematic review. PLoS Negl Trop Dis. 2022;16:e0010540. 10.1371/journal.pntd.0010540.

2. World Health Organization. Ending the neglect to attain the sustainable development goals: One health: approach for action against neglected tropical diseases 2021-2030. In World Health Organization; 2022. Available from: https://iris.who.int/handle/10665/351193. (Accessed June 1, 2025).

3. Ministry of Health. Report on the Situation of Cystic Echinococcosis/Hydatidosis in Chile, 2015– 2019. 2021. Available from: https://diprece.minsal.cl/wp-content/uploads/2021/03/Informe-situacion-de-la-Equinococosis-quistica-hidatidosis-en-chile-2015-2019.pdf. (Accessed June 5, 2025).

4. Elmoghazy W, Alqahtani J, Kim SW, Sulieman I, Elaffandi A, Khalaf H. Comparative analysis of surgical management approaches for hydatid liver cysts: conventional vs. minimally invasive techniques. Langenbecks Arch Surg. 2023;408:320. 10.1007/s00423-023-03043-8.

5. Abbasi Dezfouli S, El Rafidi A, Aminizadeh E, Ramouz A, Al-Saeedi M, Khajeh E, et al. Risk factors and management of biliary leakage after Endocystectomy for hepatic cystic echinococcosis. PLoS Negl Trop Dis. 2023;17:e0011724. 10.1371/journal.pntd.0011724.

6. Al-Saeedi M, Khajeh E, Hoffmann K, Ghamarnejad O, Stojkovic M, Weber TF, et al. Standardized endocystectomy technique for surgical treatment of uncomplicated hepatic cystic echinococcosis. PLoS Negl Trop Dis. 2019;13:e0007516. 10.1371/journal.pntd.0007516.

7. Ibrahim I, Yasheng A, Tuerxun K, Xu Q-L, Tuerdi M, Wu Y-Q. Effectiveness of a Clinical Pathway for Hepatic Cystic Echinococcosis Surgery in Kashi Prefecture, Northwestern China: A Propensity Score Matching Analysis. Infect Dis Ther. 2021;10:1465–77. 10.1007/s40121-021-00466-y.

8. Shi H, Tuerxun K, Yusupu A, Yusupu Z, Xu Q, Jia Y, et al. Perioperative outcomes and hospitalization costs of radical vs. conservative surgery for hepatic cystic echinococcosis: A retrospective study. PLoS Negl Trop Dis. 2024;18:e0012620. 10.1371/journal.pntd.0012620.

9. Huang L, Zheng B, Aduo, Ouzhulamu, Li X, Yao J. Association between radical versus conservative surgery and short-term outcomes of hepatic cystic echinococcosis in Nyingchi, China: a retrospective cohort study. BMC Surg. 2023;23:126. 10.1186/s12893-023-02000-y.

10. Baimakhanov Z, Kaniyev S, Serikuly E, Doskhanov M, Askeyev B, Baiguissova D, et al. Radical versus conservative surgical management for liver hydatid cysts: A single-center prospective cohort study. JGH Open. 2021;5:1179–82. 10.1002/jgh3.12649.

11. Ramia JM, Serrablo A, Serradilla M, Lopez-Marcano A, De La Plaza R, Palomares A. Major hepatectomies in liver cystic echinococcosis: A bi-centric experience. Retrospective cohort study. Int J Surg. 2018;54:182–6. 10.1016/j.ijsu.2018.04.049.

12. McIsaac DI, MacDonald DB, Aucoin SD. Frailty for perioperative clinicians: a narrative review. Anesth Analg. 2020; 130:1450–1460. 10.1213/ANE.0000000000004602.

13. Cappe M, Laterre PF, Dechamps M. Preoperative frailty screening, assessment and management. Curr Opin Anesthesiol. 2023;36:83–8. 10.1097/ACO.0000000000001221.

14. De Biasio JC, Mittel AM, Mueller AL, Ferrante LE, Kim DH, Shaefi S. Frailty in critical care medicine: a review. Anesth Analg. 2020;130:1462–73. 10.1213/ANE.0000000000004665.

15. Farhat JS, Velanovich V, Falvo AJ, Horst HM, Swartz A, Patton Jr JH, Rubinfeld IS. Are the frail destined to fail? Frailty index as predictor of surgical morbidity and mortality in the elderly. J Trauma Acute Care Surg. 2012;72:1526–30. 10.1097/TA.0b013e3182542fab.

16. Velanovich V, Antoine H, Swartz A, Peters D, Rubinfeld I. Accumulating deficits model of frailty and postoperative mortality and morbidity: its application to a national database. J Surg Res. 2013;183:104–10. 10.1016/j.jss.2013.01.021.

17. Subramaniam S, Aalberg JJ, Soriano RP, Divino CM. New 5-factor modified frailty index using American College of Surgeons NSQIP data. J Am Coll Surg. 2018;226:173e181.e8. 10.1016/j.jamcollsurg.2017.11.005.

18. Mouchtouris N, Luck T, Locke K, Hines K, Franco D, Yudkoff C, et al. Comparison of 5-Item and 11-Item Modified Frailty Index as Predictors of Functional Independence in Patients With Spinal Cord Injury. Global Spine J. 2025;15:782–9. 10.1177/21925682231211279.

19. Morley JE, Malmstrom TK, Miller DK. A simple frailty questionnaire (FRAIL) predicts outcomes in middle aged African Americans. J Nutr Health Aging. 2012;16:601–8. 10.1007/s12603-012-0084-2.

20. Raîche M., Hébert R., Dubois M. PRISMA-7: A case-finding tool to identify older adults with moderate to severe disabilities. Arch Gerontol Geriatr. 2008;47:9–18. 10.1016/j.archger.2007.06.004.

21. Vandenbroucke JP, von Elm E, Altman DG, Gøtzsche PC, Mulrow CD, Pocock SJ, et al; STROBE Initiative. Strengthening the Reporting of Observational Studies in Epidemiology (STROBE): explanation and elaboration. Int J Surg. 2014;12:1500–24. 10.1016/j.ijsu.2014.07.014.

22. Clavien PA, Barkun J, de Oliveira ML, Vauthey JN, Dindo D, Schulick RD, et al. The Clavien-Dindo classification of surgical complications: five-year experience. Ann Surg. 2009;250:187–96. 10.1097/SLA.0b013e3181b13ca2.

23. WHO Informal Working Group. International classification of ultrasound images in cystic echinococcosis for application in clinical and field epidemiological settings. Acta Trop. 2003;85:253–61. 10.1016/s0001-706x(02)00223-1.

24. Collado-Aliaga J, Romero-Alegría A, Alonso-Sardón M, Muro A, López-Bernus A, Velasco-Tirado V, et al. Complications Associated with Initial Clinical Presentation of Cystic Echinococcosis: A 20-year Cohort Analysis. Am J Trop Med Hyg. 2019;101:628–35. 10.4269/ajtmh.19-0019.

25. Mejri A, Arfaoui K, Omry A, Yaakoubi J, Mseddi MA, Rchidi J, et al. Acute intraperitoneal rupture of hydatid cysts of the liver: Case series. Medicine (Baltimore). 2021;100:e27552. 10.1097/MD.0000000000027552.

26. Manterola C, Vial M, Sanhueza A, Contreras J. Intrabiliary rupture of hepatic echinococcosis, a risk factor for developing postoperative morbidity: a cohort study. World J Surg. 2010;34:581–6. 10.1007/s00268-009-0322-x.

27. Manterola C, Rivadeneira J, Otzen T, Rojas-Pincheira C. Cholangiohydatidosis. Clinical features, postoperative complications and hospital mortality. A systematic review. PLoS Negl Trop Dis. 2024;18:e0011558. 10.1371/journal.pntd.0011558.

28. Castillo S, Manterola C, Grande L, Rojas C. Infected hepatic echinococcosis. Clinical, therapeutic, and prognostic aspects. A systematic review. Ann Hepatol. 2021;22:100237. 10.1016/j.aohep.2020.07.009.

29. Manterola C, Rivadeneira J, Otzen T, Rojas-Pincheira C. Hepato-thoracic cystic echinococcosis transit. Clinical features, postoperative complications and hospital mortality. A systematic review. HPB. 2025;27:330–42. 10.1016/j.hpb.2024.12.001.

30. American Society of Anesthesiologists. Statement on ASA physical status classification system. Available online: https://www.asahq.org/standards-and-practice-parameters/statement-on-asa-physical-status-classification-system (Accessed June 15, 2025).

31. Zhang Y, Wu Q, Han M, Yang C, Kang F, Li J, Hu C, Chen X. Frailty is a Risk Factor for Postoperative Complications in Older Adults with Lumbar Degenerative Disease: A Prospective Cohort Study. Clin Interv Aging. 2024;19:1117–26. 10.2147/CIA.S462731.

32. Savalei V. What to Do About Zero Frequency Cells When Estimating Polychoric Correlations. Structural Equation Modeling. 2011;18:253–73. 10.1080/10705511.2011.557339.

33. DeLong ER, DeLong DM, Clarke-Pearson DL. Comparing the areas under two or more correlated receiver operating characteristic curves: a nonparametric approach. Biometrics. 1988;44:837–45.

34. World Medical Association. Manual for physicians about the role of ethics in medicine. Available at: https://www.wma.net/what-we-do/education/medical-ethics-manual. (Accessed June 21, 2025).

35. Alkadri J, Hage D, Nickerson LH, Scott LR, Shaw JF, Aucoin SD, McIsaac DI. A systematic review and meta-analysis of preoperative frailty instruments derived from electronic health data. Anesth Analg. 2021; 133:1094–1106. 10.1213/ANE.0000000000005595.

36. Clements NA, Gaskins JT, Martin RCG 2^nd^. Predictive Ability of Comorbidity Indices for Surgical Morbidity and Mortality: a Systematic Review and Meta-analysis. J Gastrointest Surg. 2023;27:1971–87. 10.1007/s11605-023-05743-4.

37. Yalzadeh D, Cho NY, Tabibian D, Song J, Cherif A, Badiee B, et al. Comparison of frailty measures in predicting outcomes after emergency general surgery. Surgery. 2025;182:109317. 10.1016/j.surg.2025.109317.

38. Chong E, Ho E, Baldevarona-Llego J, Chan M, Wu L, Tay L. Frailty and Risk of Adverse Outcomes in Hospitalized Older Adults: A Comparison of Different Frailty Measures. J Am Med Dir Assoc. 2017;18:638.e7–638.e11. 10.1016/j.jamda.2017.04.011.

39. Chong E, Ho E, Baldevarona-Llego J, Chan M, Wu L, Tay L, et al. Frailty in Hospitalized Older Adults: Comparing Different Frailty Measures in Predicting Short- and Long-term Patient Outcomes. J Am Med Dir Assoc. 2018;19:450–457.e3. 10.1016/j.jamda.2017.10.006.

40. Konstantinidis IT, Lewis A, Lee B, Warner SG, Woo Y, Singh G, et al. Minimally invasive distal pancreatectomy: greatest benefit for the frail. Surg Endosc. 2017;31:5234–40. 10.1007/s00464-017-5593-y.

41. Loecker C, Schmaderer M, Zimmerman L. Frailty in Young and Middle-Aged Adults: An Integrative Review. J Frailty Aging. 2021;10:327–33. 10.14283/jfa.2021.14.

42. Patel I, Hall LA, Osei-Bordom D, Hodson J, Bartlett D, Chatzizacharias N, et al. Risk factors for failure to rescue after hepatectomy in a high-volume UK tertiary referral center. Surgery. 2024;175:1329–36. 10.1016/j.surg.2024.01.025.

